# Implementation of nursing process in Ethiopia and its association with working environment and knowledge: a systematic review and meta-analysis

**DOI:** 10.1101/19008144

**Authors:** Wondimeneh Shibabaw, Yared Asmare, Tadesse Yirga, Abate Dargie

## Abstract

**Background:** Nursing Process is a scientific problem solving approach to direct nurses in caring for clients effectively and to improve quality of health care service. In Ethiopia, the national pooled prevalence of implementation of nursing process remains unknown. Hence, the objective of this systematic review and meta-analysis was to estimate the level of implementation of nursing process and it association with knowledge and working environment.

**Methods:** PubMed, Scopus, Cochrane Library, Google Scholar, PsycINFO and CINAHL were systematically searched online to retrieve related articles. The Preferred Reporting Items for Systematic Review and Meta-Analysis (PRISMA) guideline was followed. To investigate heterogeneity across the included studies, I^2^ test was employed. Publication bias was examined using funnel plot and Egger’s regression test statistic. The random-effect model was fitted to estimate the summary effects, and odds ratios (ORs). All statistical analysis was done using STATA version 14 software for windows.

**Results:** Seven studies which comprises of 1,268 participants were included in this meta-analysis. The estimated pooled prevalence of implementation of nursing process in Ethiopia was 42.44% (95% CI (36.91, 47.97%)). Based on the subgroup analysis, the highest implementation of nursing process was observed sample size greater than or equals to two hundred, 44.69% (95% CI: 35.34, 54.04). Nurses who have been work in the stressful environment [(OR 0.41, 95%CI (0.08, 2.12)] and having good knowledge about nursing process [(OR 2.44, 95%CI (0.34,17.34)] was not significant associated with the implementation of nursing process.

**Conclusion:** The overall implementation of nursing process in Ethiopia is relatively low. Nurses who have been work in the stressful environment have less likely implement nursing process. On the other hand, Nurses who had good knowledge on nursing process were more likely to implement nursing process. Therefore, policymakers (FMOH) and other concerned body need give special attention to improve the implementation of nursing process.

## Background

Nursing is an emerging profession with a unique perspective on people, environment and health. Now a day the approach of client care is moved from the medical model to a holistic care[1]. A long with it nursing process play pivotal role through patient centred, goal oriented, way to improve the quality of nursing care, and to meet the individualized health care needs of their clients [2–6].

Nursing process has providing holistic patient care and serve as a key for nursing practice and education [7]. Standard implementation of nursing process could improve quality of care and encourages the utilization of evidence based nursing practice [8, 9]. Appropriately, implementation of nursing process, would provide meaning and relevance to professional knowledge [10]. Globally, nursing process is an integral part of nursing education, practice, dynamic client care and a tool for critical thinking in order to address the need of the clients [11].

Nursing process is a vital structure that provides order and direction to nursing care. Likewise, it is the corner stone of the nursing profession [12, 13]. Using nursing care plan as a tool to guide nursing care will allow nurses to make some independent decisions that can encourage healing [14]. Even though, nursing process has become the framework for nursing care, still the perception persists that it is time consuming and impractical [15].

The utilization of nursing process could assure nurses accountability and responsibility for the patient care and enable to measure quality of nursing care [16]. The essence of nursing process lies on the great benefit to the client and nursing profession [17]. In addition, nursing process guide the nursing activities in a better way, promotes quality of care, and provides professional autonomy[18]. The implementation of nursing process has been reported in different literature, across different region of the globe. For example, the study done in Kenya shows 33.1% [6], 25% in Brazil [8], and 57.1% in Nigeria [1], 81.77% in Brazil [19]. On the contrary, a study done in Congo shows about 100% [20] was not implementing nursing process.

The implementation of nursing process is complex phenomenon and due to the presence of multiple impeding factors. Identifying associated factors used as benchmarks to design appropriate measure, to improve client safety and efficient utilization of resources. Several associated factors are responsible for reducing the implementation of nursing process. For instance, lack of theoretical and practical knowledge [20–26], institutional factors[6, 11], professional factors[11],work overload [11, 21, 23, 25–28], lack of previous experience [6, 27], inadequate nursing staff [20], patients with complicated case[24], working in a hospital[25], patient economic status[25], early discharge [25, 26], not cooperative [25], lack of training [6, 18, 22, 26, 29], low educational status[27, 28], poor Nurses skill [28], lack of time [18, 20, 27, 30], lack of authorities’ support [27] and lack of equipment and material [20, 27, 28], were some of the factors associated with implementation of nursing process. On the contrary, a study conducted in Nigeria shows that institutional factor does not pose a barrier to the use of nursing process [30, 31].

Despite, the effort of Ethiopian Federal Ministry of Health to prepared and distributed standardized nursing care plan for all health care settings since 2011[32]. The implementation of nursing process in different health care setting is not well developed and organized [24, 30, 33]. However, currently many health care settings demand the implementation of nursing process in clinical practice.

In Ethiopia, Nurses constitute the backbone of healthcare delivery system to improve the quality of health care service and implementation of nursing process may contribute a significant role. Different studies have been conducted to determine the implementation of nursing process and its associated factors. The findings of these fragmented studies revealed that there was a great variability in the implementation of nursing process across the country. Hence, this study aimed to estimate the pooled prevalence of implementation of nursing process in Ethiopia and its association with working environment and knowledge. Finding from the current study would serve as input for educators, clinicians, program planner, and policy makers working in the area of nursing process implementation to design a new approach or strengthen the existing nursing process.

## Methods

### Design and Search strategy

A two-step search strategy was used to identify all relevant literature. First, six electronic databases such PubMed, Cochrane Library, Google Scholar, CINAHL, Scopus, and PsycINFO were searched to extract all available literature. Second, a hand search of gray literature and other related articles in order to identify additional relevant research. In addition, all electronic sources of information were searched from conception to April 1^st^, / 2019. The search strategy was developed using Population Exposure Controls Outcome and study design (PECOS) searching guide. The search was conducted using the following MeSH and free-text terms: “nursing process”, “implementation”, “nursing process implementation”, and “Ethiopia”. Boolean operators like “AND” and “OR” were used to combine search terms.

### PECO guide

#### Population

All Nurses working in different health care setting with experience of greater than or equal to six months.

#### Exposure

Nurses who have good knowledge on nursing process, and working on well-organized environment.

#### Comparison

Nurses who have poor knowledge on nursing process and working on stressful environment.

#### Outcome

Implementation of nursing process

### Eligibility criteria

#### Inclusion criteria

Studies were included if they met the following criteria: (1) those articles conducted in Ethiopia;(2) articles published in peer reviewed journals and gray literature;(3) published in the English language from inception to 2019; (4) report outcome variable as implementation of nursing process and if they report associated factors like working environment and knowledge on nursing process were considered.

#### Exclusion criteria

Studies were excluded on any one of the following conditions: (1) articles which didn’t fully accessed at the time of our search process ;(2) studies with poor quality score as per stated criteria ;(3) articles in which fail to determine the outcome(implementation of nursing process). Finally two authors (W.S. and Y.A.) independently evaluated the eligibility of all retrieved studies, and any disagreement and inconsistencies were resolved by discussion and consensus, with other third authors.

### Outcome of interest

The outcome of this study was the overall implementation of nursing process. The implementation of nursing process was regularly accepted from patients’ records or from verbal reports of nurses working in a hospital or outpatient unit, in at least one of the following phases: data collection, nursing diagnosis, prescription of nursing and evaluation of nursing [22, 25, 33]. The determinant variables included in this review were; working environment (well-organized vs stressful) and knowledge on nursing process (good knowledge vs poor knowledge).

### Data extraction and quality assessment

Data were extracted by two authors using a pre-piloted and standardized data extraction format prepared in a Microsoft Excel spread sheet. Data on author/s name, year of publication, study area/Region, health institution, study design, sample size, prevalence, determinant factors and the quality score of each study were extracted from each included article by three independent authors (Y.A,W.S, and T.Y). The quality of each study was assessed using modified version of the Newcastle-Ottawa Scale for cross-sectional study[34]. Studies were included in the analysis if they scored ≥5 out of 10 points in three domains of modified NOS components for cross-sectional studies [34, 35]. The score point of each domain is selection (5 points), comparability (2 points), and outcome assessment (3 points). Finally, the quality score of each study was extracted from incorporated article by three independent authors. Any disagreements at the time of data abstraction were resolved by discussion and consensus.

Supplementary file 1: Methodological quality assessment of cross-sectional studies using modified Newcastle - Ottawa Scale (NOS).

### Assessment of risk of bias in included studies

The risk of bias tool for prevalence studies developed by Hoy and colleagues [36] was used to assess the risk of bias among included studies reporting on the prevalence of implementation of nursing process (supplementary file 2 for Hoy et al tool). On the other hand, the Quality in Prognosis Studies tool was used to assess the risk of bias for studies reporting the factors associated with the implementation of nursing process [37]. Both authors were undertake the risk of bias assessment of the included studies independently.

### Statistical analysis

A meta-analysis of the implementation of nursing process was carried out using a random-effects (DerSimonian and Laird) method since it is the most common method in a meta-analysis to adjust for the observed variability [38]. Heterogeneity across the studies was checked using the I^2^ statistics test[38]. In addition, to investigate the possible sources of heterogeneity, meta-regression and subgroup analysis were deployed. Publication bias was assessed by visual inspection of a funnel plot. Similarly, Egger test was conducted and a p□≤□0.05was considered statistically significant for the presence of publication bias [39, 40]. Moreover, sensitivity analysis was performed to investigate whether the pooled effect size was influenced by individual studies. The pooled effect size (i.e. proportion and odds ratio (OR)) with a 95% confidence interval (CI) was generated and presented using a forest plot. All data manipulation and statistical analyses were performed using Stata version 14.0 software for Windows [41].

### Presentation and reporting of results

The Preferred Reporting Items for Systematic Reviews and Meta-analyses (PRISMA) guideline was used to publish the proposed systematic review[42]. The PRISMA checklist was used alongside the final review (supplementary file 3). The entire process of study screening, selection and inclusion were depicted with the aid of a flow diagram. A reason for study exclusion was documented and summary shown in the flow diagram. Quantitative data were presented on forest plots and summary tables.

## Result

### Search results

We found that a total of 648 articles, of these, 643 studies were found from six international databases and the remaining 5 were through manual search. Databases includes; PubMed (4), Scopus (83), psyInfo (46), Cochrane library data base (68), Google scholar (327), and CINHAL (115). Out of them, 239 duplicate records were identified and removed. From the remaining 409 articles, 363 articles were excluded after reading of titles and abstracts based on the pre-defined inclusion criteria’s. Finally, 29 full text articles were assessed for eligibility criteria. Based on the pre-defined criteria and quality assessment, only 7 articles were included for the final analysis (figure 1).

**Figure 1.**
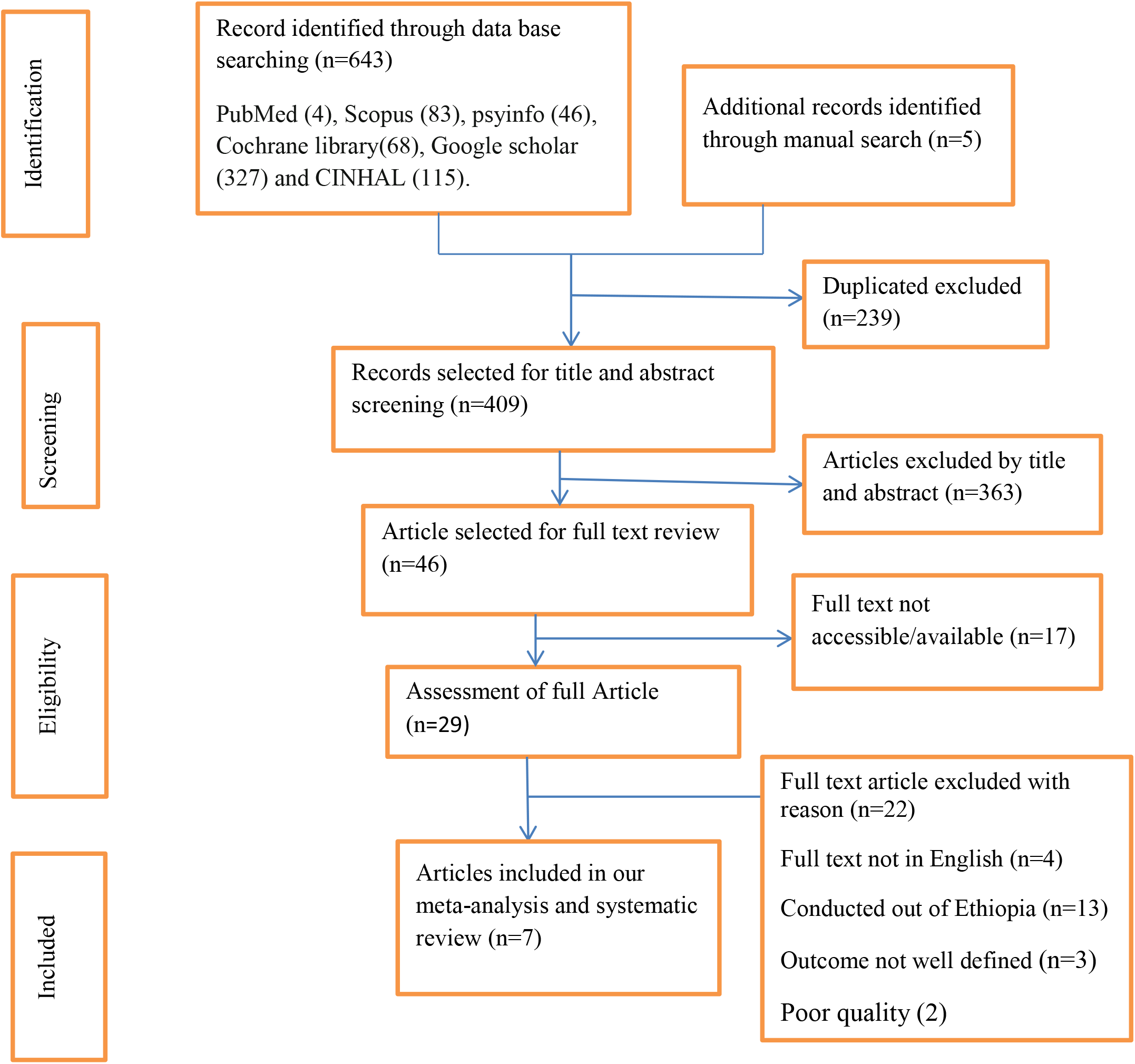
PIRSMA Flowchart diagram of the study selection.

### Baseline characteristic of the included studies

A total of 7 studies with 1,268 participants were included in this meta-analysis. Of those, two studies [24, 33] were conducted in Amharic region, one [22] in Afar region, one [23] in Addis Ababa, one [29] in Harari region, and the remaining one [28] in Tigray region. Regarding sample size more than half of the studies (57.14%) were less than two hundred [22, 24–26]. The highest prevalence of implementation of nursing process (52.1%) was reported from Addis Ababa and the lowest (32.7%) in South Nation Nationality and People. Based on modified Newcastle Ottawa score quality assessment almost all seven articles fulfil the required quality. All the included studies were cross-sectional by design and were conducted among nurses working in different clinical setting of Ethiopia (Table1).

### Implementation of nursing process in Ethiopia

The result of this meta-analysis using random effects model showed that the pooled prevalence of implementation nursing process in Ethiopia was 42.44% (95% CI: 36.90-47.97) (Figure 2) with high significant level of heterogeneity was observed (I^2^ = 74.1%; p < 0.001). The presence of significant magnitude of heterogeneity also suggests the need to conduct subgroup analysis. To identifying the sources of heterogeneity, there is a need to employed sub group analysis by using study sample size to evaluate the implementation nursing process (Figure 3). The finding of subgroup analysis using sample size showed that the highest implementation of nursing process was observed studies done using sample size was greater than or equals to 200 (44.7% (95% CI: 35.34,54.04), I^2^ = 84.8%).

**Figure 2.**
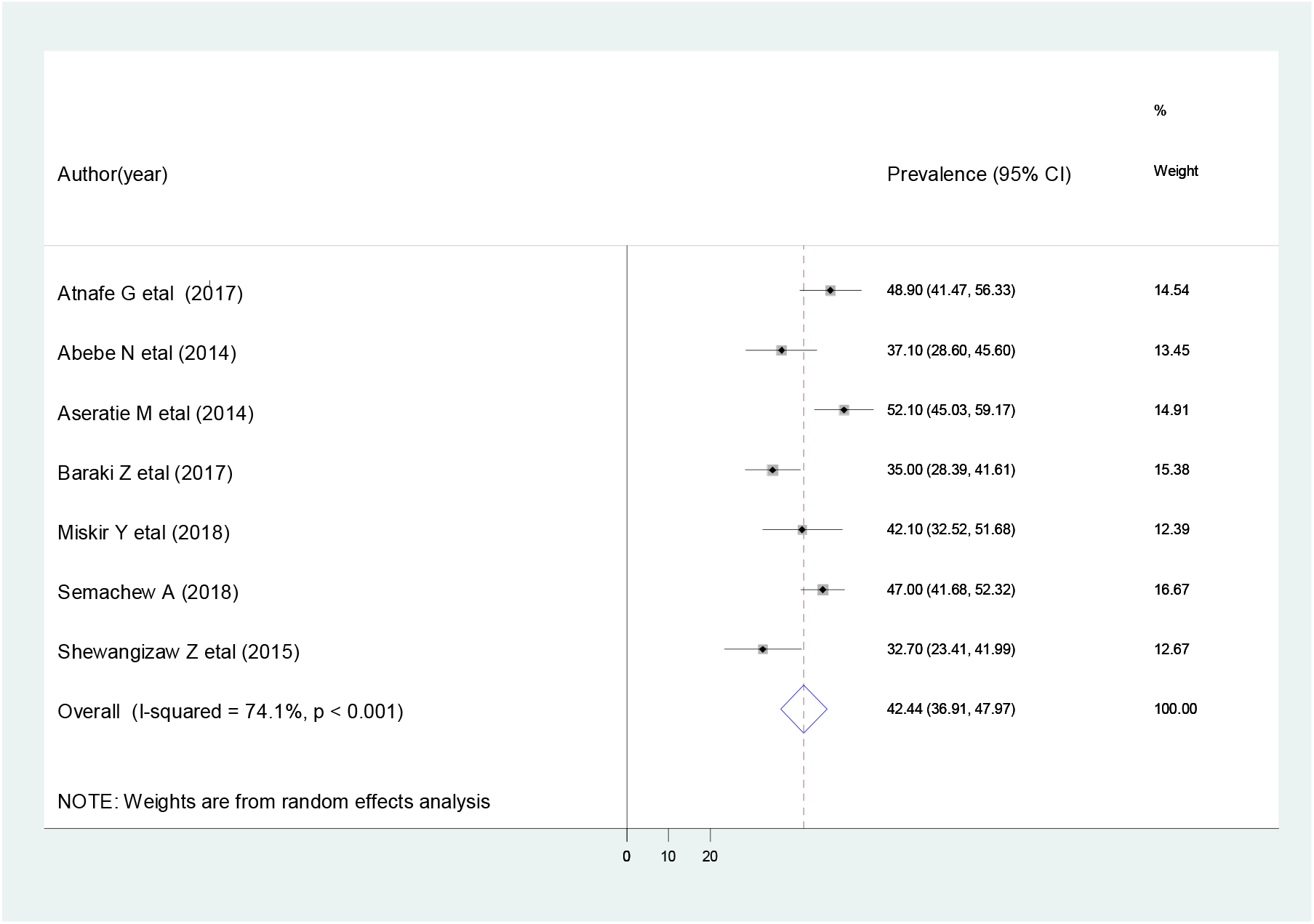
Forest plot showing the pooled prevalence of implementation nursing process.

**Figure 3.**
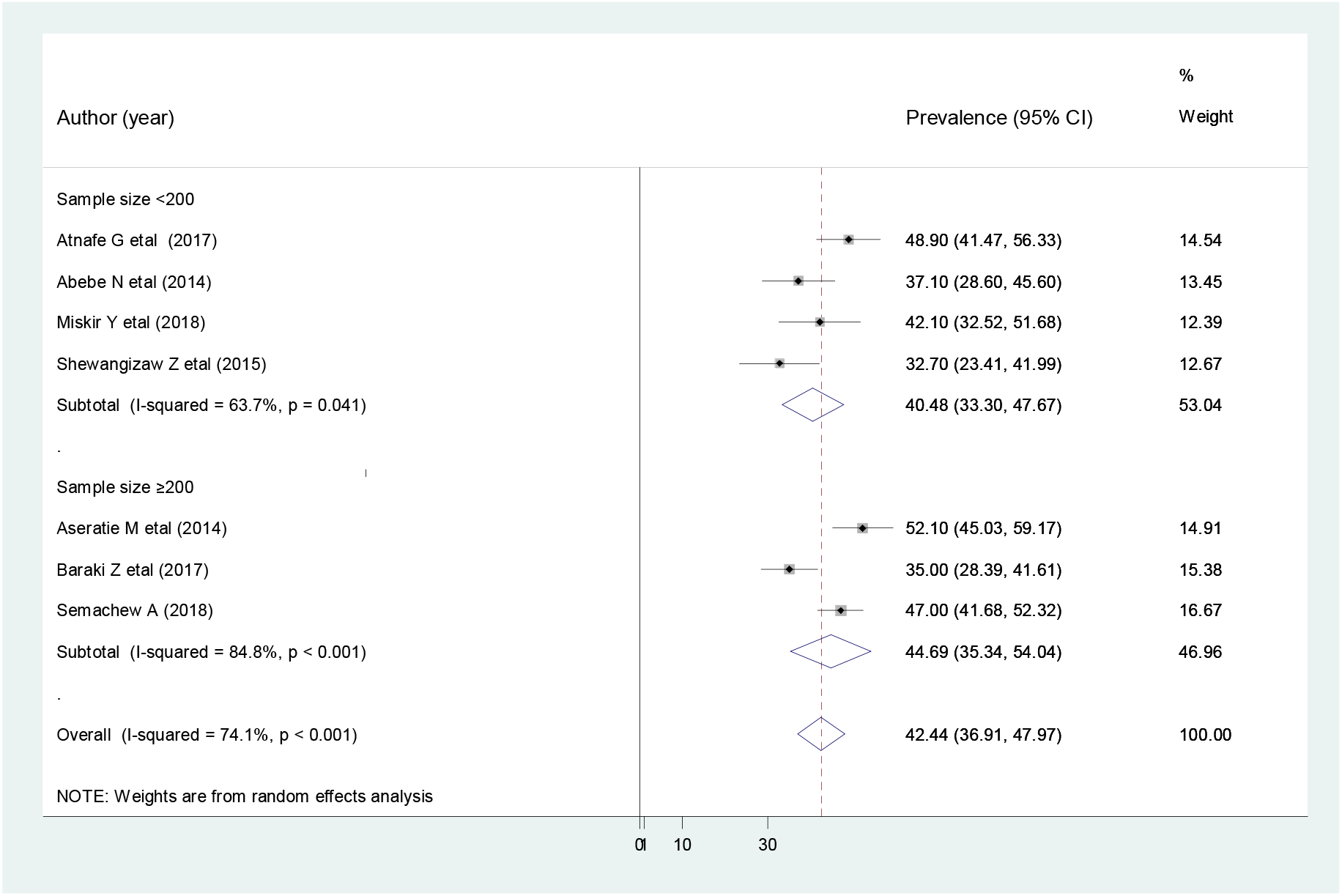
Subgroup analysis by sample size on the implementation nursing process.

### Investigating source of heterogeneity

As the test statistic shows that there is a significant heterogeneity within and between the included studies (I^2^ statistics=74.1%). Hence, to minimize the source of heterogeneity between the point estimates of the primary study, subgroup analysis was done based on study sample size. In addition, in order to identify the possible source of variation across the included studies, we have performed meta-regression by using publication year and sample size of each article as covariate of interest. However, the result of the meta-regression analysis showed that both covariates were not statistically significant for the presence of heterogeneity (Table 2).

**Table 1:** Baseline characteristics of studies included in the meta-analysis.

| Primary Author | Pub. Year | Study area, Region | Health Facility Name | Study Design | Sample Size | Prevalence % (95% CI) | Quality score |
| --- | --- | --- | --- | --- | --- | --- | --- |
| Abebe N etal [24] | 2014 | Amhara | Finoteselam &debre markos hospital | Cross-sectional | 139 | 37.1(28.6-45.6) | 8 |
| Miskir Y etal [22] | 2018 | Afar | Afar region hospitals | Cross-sectional | 107 | 42.1(32.5- 51.6) | 7 |
| Aseratie M etal [23] | 2014 | Addis ababa | Public hospitals | Cross-sectional | 202 | 52.1(45.0-59.2) | 8 |
| Shewangizaw Z etal [25] | 2015 | Arba Minch, SNNP | Arbaminch General Hospital | Cross-sectional | 105 | 32.7(23.4- 41.9) | 8 |
| Baraki Z etal [28] | 2017 | Tigray | hospitals of Central and Northwest zones, | Cross-sectional | 200 | 35.0(28.4- 41.6) | 7 |
| Semachew Ayele [33] | 2018 | Amhara | Felege Hiwot Referral hospital, Debretabor and Finoteselam general hospitals | Cross-sectional | 338 | 47.0(41.7- 52.3) | 7 |
| Atnafe G etal [29] | 2017 | Harari | Public Hospitals of Harari People National Regional State | Cross-sectional | 177 | 48.9(41.5-56.3) | 6 |

**Table 2.** Meta regression analysis for the included studies to identify source of heterogeneity.

| Covariate(source) | Coefficients | Standard error | p-value | 95%CI |
| --- | --- | --- | --- | --- |
| Publication year | -0.0102555 | 1.011351 | 0.992 | -6.062, 5.131 |
| Sample size | 0.0473206 | 0.0434728 | 0.338 | -0.0733,0.168 |

### Publication bias

To identify the presence of publication bias, funnel plot, and egger’s test was performed. The visual inspection of the funnel plots showed symmetrical distribution, which is the evidence for absence publication bias (Figure 4). Likewise, symmetry of the funnel plot was not statistically significant as evidenced by egger test (P=0.349). Furthermore, the findings of sensitivity analyses using random effects model revealed that no single study affect the overall implementation of nursing process (figure 5).

**Figure 4:**
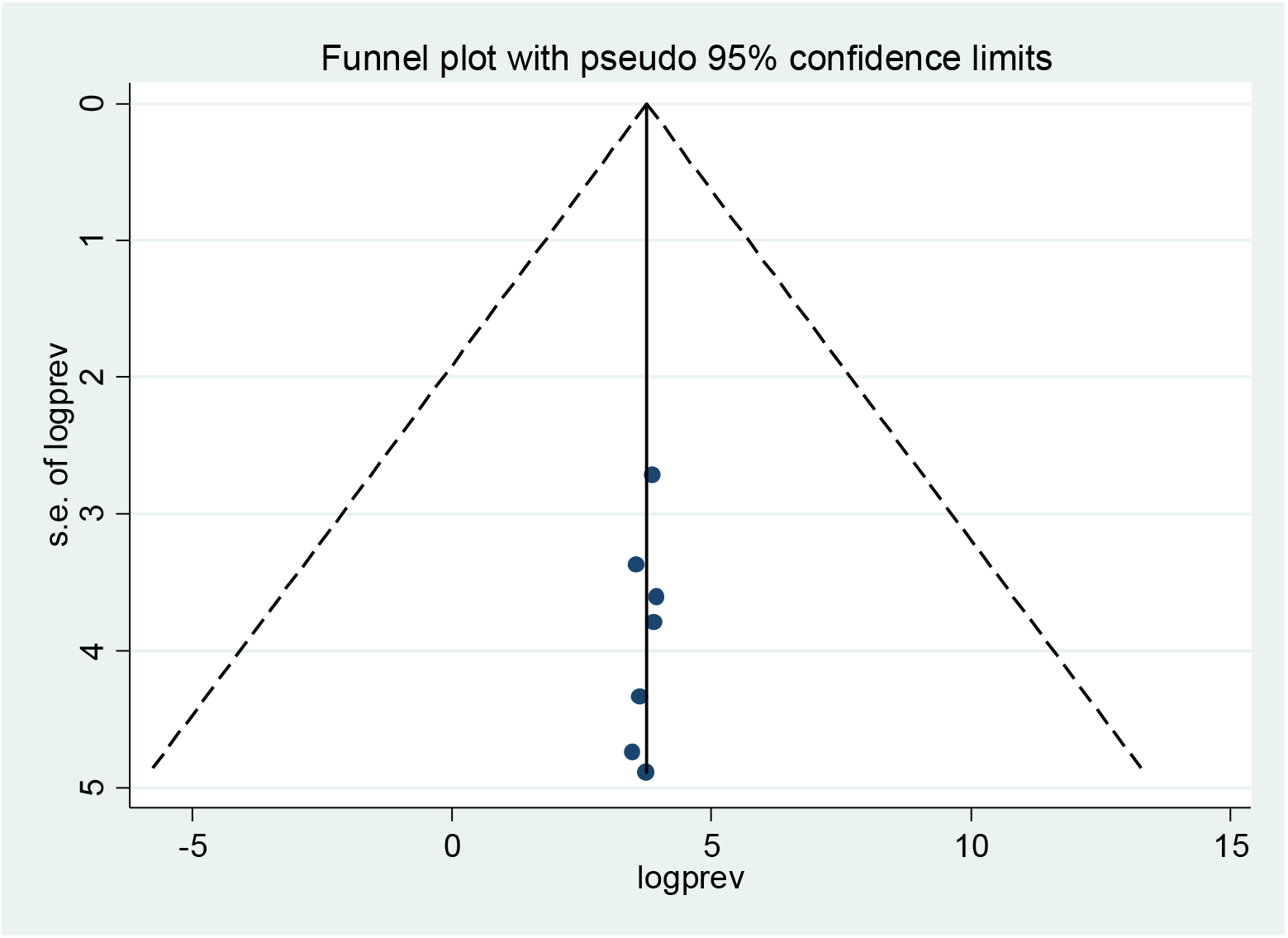
Funnel plot to test the presence of publication bias of the 7 studies.

**Figure 5:**
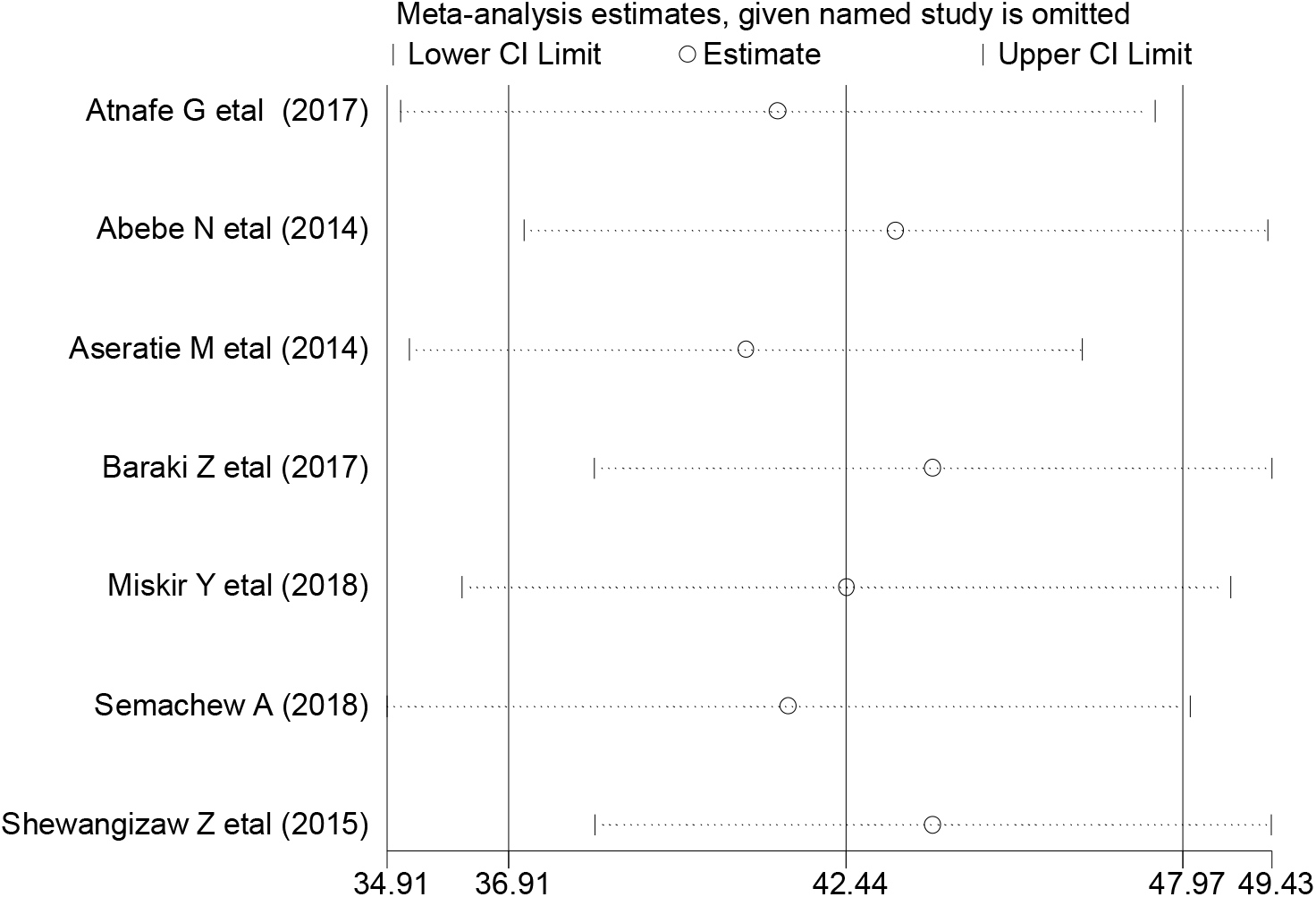
Result of sensitivity analysis of the 7 studies.

**Figure 6:**
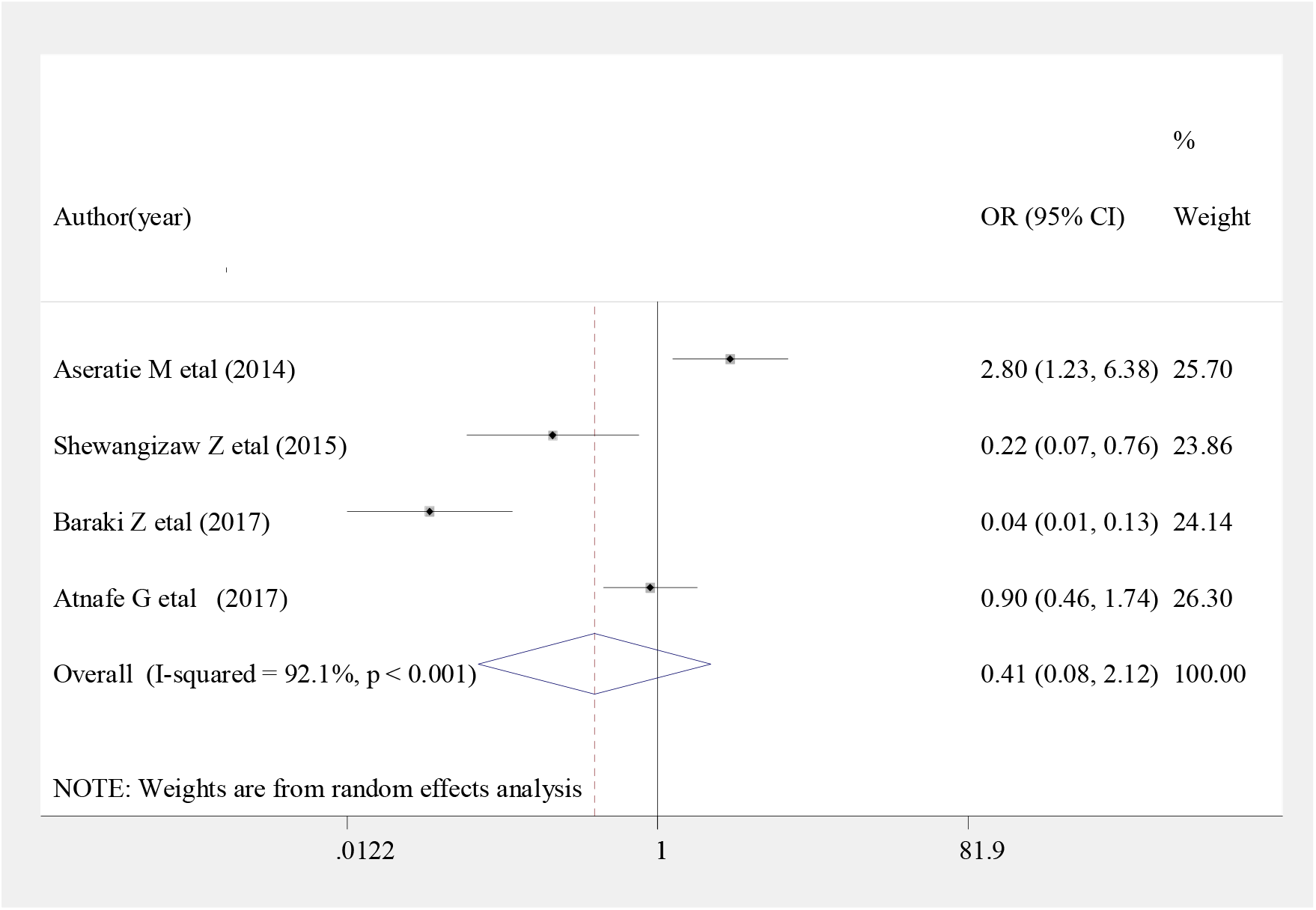
Forest plot showing the association between implementation nursing process and nurse working environment.

### Association between working environment and implementation of nursing process

The finding of the current meta-analysis revealed that, Nurses who are working in the stressful area were 59% less likely to implement nursing process compared to those working in a well-organized area, (OR: 0.41 (95% CI (0.08, 2.12)) (Figure 5), even if not statistically significant. The heterogeneity test (P= 0.000) shows significant evidence of variation across studies. Moreover, the evidence from Egger’s test shows that there was a no significant proof of publication bias (P = 0.291).

### Association between knowledge and implementation of nursing process

According to the finding of recent meta-analysis study, Nurses who had good knowledge on nursing process had 2.44 times higher chance of being implementing nursing process compared to those who have poor knowledge (OR: 2.44 (95% CI (0.34,17.34)) (Figure 7). Though, the finding is not statistically significant. The heterogeneity test (P= 0.000) showed a significant evidence of variation across studies. However, the result of Egger’s test showed that no statistically significant evidence of publication bias (P = 0.510).

**Figure 7:**
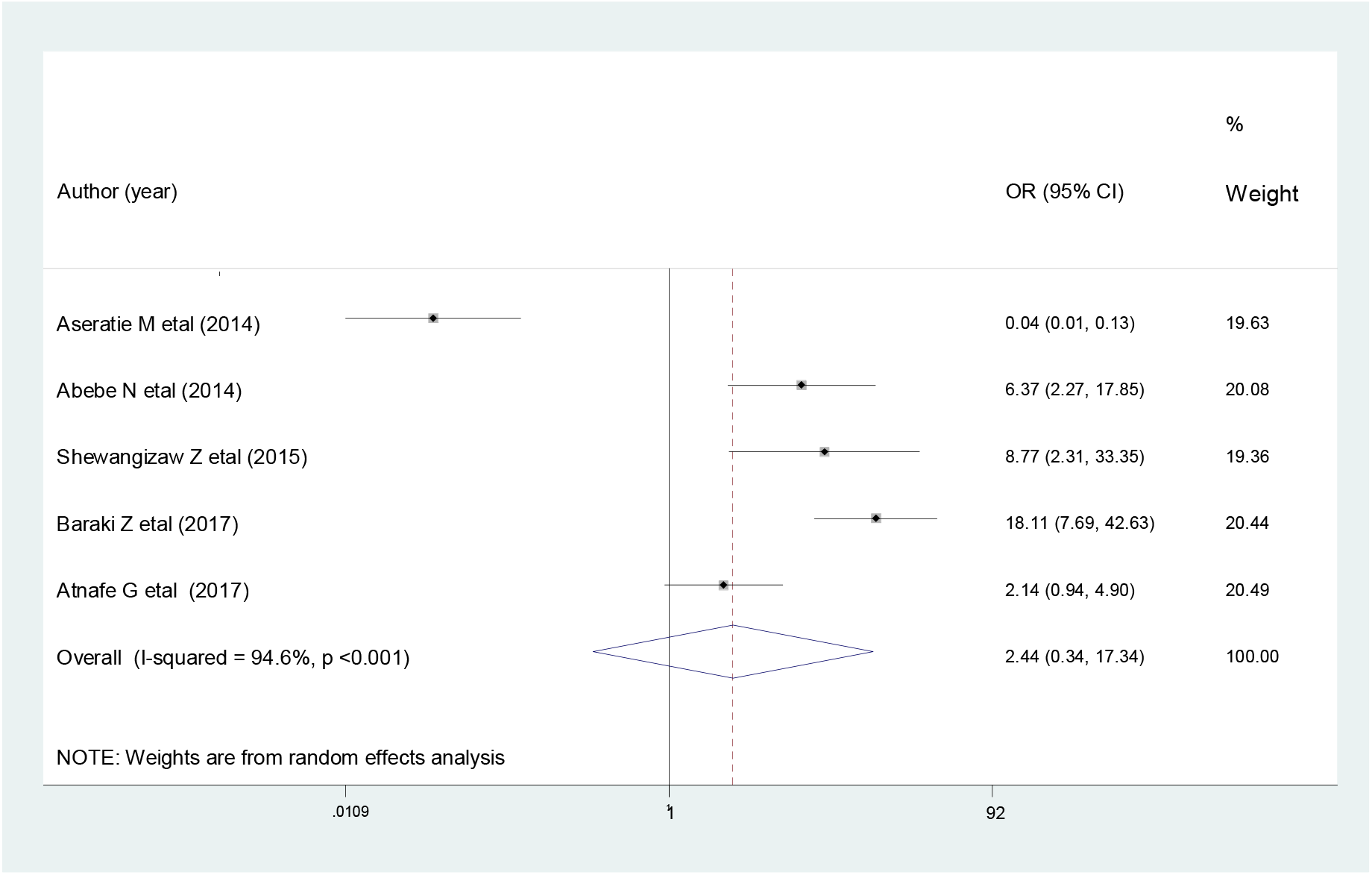
Forest plot shows the association between implementation of nursing process and nurse knowledge on nursing process.

## Discussion

The main objective of this systematic review and meta-analysis is to estimate the pooled prevalence of implementation of nursing process in Ethiopia and its association with working environment and knowledge. In line with this objective the findings of 7 included studies were revealed that the overall implementation of nursing process in Ethiopia was 42.44% (95% CI: 36.9, 47.9%). The finding of the current study indicated in this meta-analysis is higher than the study done in Kenya which showed 33.1% [6], and Brazil 25% [8]. On the contrary, the finding is lower than a study done in Nigeria 57.1%[1] and 81.77% in Brazil [19]. The possible explanations for the above variations might be methodological differences (i.e., data analysis and eligibility of study participants), variation in awareness, knowledge and educational background among nurses, policy and strategy difference and health service utilization.

The result of the subgroup analysis based on sample size (i.e. sample size <200 and ≥200) showed that the highest pooled prevalence of implementation of nursing process was observed from studies done with sample size were greater than or equal to 200 which revealed that 44.7% (95%CI: 35.34, 54.04). As a result, the findings of the subgroup analysis revealed that variability was observed in the overall implementation of nursing process across the category of each sample size. The possible explanation for this variation might be if the sample size is increase it would provide the true estimate of the population.

The current meta-analysis was estimated the association between nurses knowledge on nursing process and implementation of nursing process. Nurses who had good knowledge were positively associated with the implementation of nursing process. Even though, it is not statistically significant. Nurses who had good knowledge were almost 2.44 times more likely to implement nursing process as compared to those who had poor knowledge. This finding is in line with the studies conducted in different region of the globe [43–46]. Likewise, the finding also supported with other study, in which nurses who have theoretical knowledge on the meaning of nursing process could successfully promoting the quality of care for the client through the implementation of nursing process [18, 47].

According to the current meta-analysis finding, Nurses who have been working at a stressful working environment were nearly 59% less likely to implement nursing process as compared to those who have working at well-organized environment. Even thought, it is not statistically significant. The finding is in agreement with study conducted in Egypt [48]. This might be reflect that the establishment of conducive environment to make effective and efficient nursing practice, viewed by previous researches as important aspects that affected nurses□ ability to conduct “good” practice.

Though this meta-analysis has provided important information and synthesise evidence regarding the implementation of nursing process, there are some limitations that need to be considered in the future research. First, only English articles were considered; Second, most of the studies included had small sample size; Third, it was challenging to synthesise some additional factors meanwhile they were not examined in a related approach across the studies; Fourth, all included studies were reported hospital-based data. Lastly, it was challenging to compare and contrast our finding with others because of lack of other published systematic review and meta-analysis on the implementation of nursing process.

### Implications for Nursing Practice

This meta-analysis has implication for clinical practice. Estimating the pooled prevalence of implementation of nursing process would provide current evidence to the utilization of nursing process, application of standardized care, to improve patient satisfaction, and to address the client demand. Which enable Hospital wards should be adequately staffed with nurses to prevent the situation of stressful working environment as a factor responsible for inconsistent use of nursing process. In addition, it facilitates to design different strategies for enhancing nurse knowledge on nursing process. Furthermore, the finding serves as alarming to health care professional to give a focus on the application of standardized care and represents a marker of quality of care.

### Conclusion and recommendations

The overall implementation of nursing process in Ethiopia is significantly low. Nurses who work in the stressful environment had a negative influence the implementation of nursing process. Nurses who had good knowledge on nursing process were more likely to implement nursing process. Therefore, policymakers (FMOH) could give special attention to the implementation of nursing process to improve the overall quality of healthcare service. Furthermore, further qualitative study is needed to explore the reason behind poor implementation of nursing process.

## Data Availability

The data analyzed during the current systematic review and meta-analysis is available from the corresponding author on reasonable request.

## Abbreviations

CI: Confidence Interval
FMOH: Federal Minister of Health
OR: Odds Ratio
PRISMA: Preferred Reporting Items for Systematic Reviews and Meta-Analyses
SNNP: Southern Nations, Nationalities, and Peoples
WHO: World Health Organization.

## Declaration

### Ethics approval and consent to participate

Not applicable.

### Consent for publication

Not applicable.

### Authors’ contributions

WS and TY developed the protocol and involved in the design, selection of study, data extraction, and statistical analysis and developing the initial drafts of the manuscript. YA, AD and TY involved in data extraction, quality assessment, statistical analysis and revising subsequent drafts. WS and YA prepared the final draft of the manuscript. All authors read and approved the final draft of the manuscript.

## Acknowledgements

We would like to thank all authors of studies included in this systematic review and meta-analysis.

